# Comparative Analysis of Biofilm Formation in Bacterial and Fungal Isolates from Contact Lens and Non-Contact Lens Associated Keratitis

**DOI:** 10.64898/2026.02.09.26345896

**Authors:** Kavin Shilpa Abraham, Sujith Sri Surya Ravi, Leela Kagithakara Vajravelu

## Abstract

Microbial keratitis is a sight-threatening corneal infection with varying etiological agents, primarily bacteria and fungi. Assessing and contrasting the virulence factors of microorganisms isolated from a non-contact lens-associated keratitis (NCLAK) and contact lens-associated keratitis (CLAK) is the goal of the current investigation. Samples were collected from over 60 patients and analysed using standard microbiological techniques, including culture, Gram staining, KOH mount, biochemical tests, antimicrobial susceptibility testing, and biofilm assays. The results demonstrated that CLAK isolates were predominantly bacterial, especially *Pseudomonas aeruginosa*, known for strong biofilm production and high multidrug resistance. In contrast, NCLAK showed a higher incidence of fungal infections, particularly *Candida albicans*. The results highlight the significance of early diagnosis, tailored and improved awareness regarding contact lens hygiene to prevent complications associated with keratitis.

## 1. Introduction

Keratitis, defined as inflammation of the cornea, is a leading cause of visual impairment and blindness worldwide, particularly in developing nations[1]. The cornea, being the transparent and avascular front portion of the eye, is constantly exposed to external environmental factors such as dust, microorganisms, trauma, and foreign bodies[2]. As such, it serves as a crucial anatomical and functional barrier against pathogens. Infections of the cornea, if left untreated or improperly managed, can result in corneal scarring, opacification, perforation, and permanent vision loss[3].

Microbial keratitis, specifically, refers to infections caused by bacteria, fungi, viruses, and protozoa, with bacterial and fungal etiologies being the most prevalent[4]. It is a significant public health issue in tropical and sub-tropical nations like India, frequently presenting in ophthalmology clinics[5].

Contact lens wear is considered a predominant risk factor in urban and semi-urban settings, especially with the rising popularity of cosmetic lenses and increased access to over-the-counter lens purchases[6]. Poor lens hygiene habits, such as leaving lenses on all night, not washing them, using them again, and using solutions, can result in contamination and colonization of the lens surface by pathogenic organisms. *P. aeruginosa (Pseudomonas aeruginosa)*, a Gram-negative bacterium with high virulence and biofilm-forming capacity, is the most prevalent infection linked to Contact Lens-associated Keratitis (CLAK)[7]. On the other hand, Non-Contact Lens-Associated Keratitis (NCLAK) primarily results from ocular trauma, often involving vegetative matter such as twigs, leaves, or soil, particularly in agricultural workers[8]. In India, filamentous fungus such as Aspergillus and Fusarium species are the most common cause of fungal keratitis, although yeast infections, such as those by Candida albicans also occur, particularly in immunocompromised individuals[9]. Unlike bacterial keratitis, fungal keratitis typically has an indolent onset, delayed diagnosis, and a prolonged treatment course, often requiring systemic antifungal therapy and, in severe cases, surgical intervention[10].

The pathogenesis of microbial keratitis involves microbial adherence to the corneal epithelium, epithelial damage, release of toxins, and stimulation of host immune responses, including the discharge of cytokines that promote inflammation[11]. Examples of virulence factors such as enzymes (proteases, phospholipases), exotoxins, and biofilms contribute significantly to the pathogenicity of both bacterial and fungal organisms. Biofilm production, in particular, plays a central role in persistent infections and antimicrobial resistance[12].

## 2. Methodology

Between August 2024 and March 2025, seven-month prospective comparative research was conducted at the SRM Medical College Hospital and Research Center, an ophthalmology outpatient department at a government hospital, and eye care clinics in the Villupuram district. The Institutional Ethics Committee approved the study, and all participants gave their informed consent.

### 2.1. Study population

A total of 60 patients who had clinical signs of microbial keratitis were included in the study. These were categorized into two groups based on history and clinical background: There are two types of keratitis: contact lens-associated keratitis (CLAK) and non-contact-lens-associated keratitis (NCLAK). Patients of both genders, aged above 18 years, were included. Exclusion criteria were recent ocular surgery, patients on topical antibiotics within 72 hours, and individuals with viral or parasitic keratitis.

### 2.2 Collection and Transportation of Sample

Samples were taken from the affected eye using sterile techniques under slit-lamp guidance. Corneal scrapings were obtained using a sterile Bard-Parker blade or platinum spatula for non-contact-lens-associated keratitis (NCLAK) cases. Additional samples included conjunctival swabs, contact lens swabs, and contact lenses flushed in saline for contact lens-associated keratitis (CLAK) cases. All collected samples were placed in sterile containers and transported immediately to the microbiology laboratory at 4°C for processing.

### 2.3 Microscopy and culture techniques

were employed for the identification of bacterial and fungal isolates. Smears were prepared and stained using Gram stain to determine bacterial morphology and to classify organisms as Gram-positive or Gram-negative. A 10% KOH mount was used for the preliminary detection of fungal elements such as hyphae, yeast cells, and pseudo hyphae, while Lactophenol Cotton Blue (LPCB) staining was performed for detailed morphological observation of fungi in culture-positive isolates. Samples were inoculated on various culture media such as MacConkey agar (MA) for the isolation of gram-negative enteric bacteria, Sabouraud Dextrose Agar (SDA) for the isolation of fungi, and nutrient agar (NA) and blood agar (BA) for the growth of general and fastidious bacteria. SDA plates underwent daily monitoring for a maximum of four weeks while being incubated at both 25°C and 37°C. Growth was considered significant if the same organism was isolated on multiple media or if heavy growth with consistent microscopic findings was observed on a single medium.

### 2.4 Biochemical identification

of isolated bacteria was performed using standard procedures. Catalase and coagulase tests were employed for the identification of *Staphylococcus* species. For Gram-negative rods, a series of biochemical tests, including oxidase, indole production, tests for urease, citrate utilization, and triple sugar iron (TSI) were performed. Colony morphology, pigmentation, and microscopic features such the presence and arrangement of Macroconidia, Microconidia, and the kind of septation were used to identify the fungal isolates.

### 2.5 Biofilm formation

The microtiter plate method was used to evaluate the isolates’ capabilities. Tryptic soy broth was used to dilute overnight cultures, which were then inoculated in 96-well flat-bottom polystyrene microtiter plates, and the plates were then incubated for 24 hours at 37°C. Following incubation, wells were gently washed with phosphate-buffered saline (PBS) to remove non-adherent cells. The remaining biofilms were stained with 0.1% crystal violet, and the bound pigment was subsequently dissolved using ethanol. The optical density (OD) at 570 nm was measured using a microplate reader. Biofilm production was classified according to OD values: OD > 0.4 denoted strong biofilm formers, OD 0.1–0.4 denoted moderate biofilm formers, and OD < 0.1 denoted non-biofilm formers.

### 2.6 Data Collection and Statistical Analysis

All the results were entered into Microsoft Excel. Descriptive statistics including mean, standard deviation, and percentages were used to summarize the data. As needed, chi-square and t-tests were employed to compare the CLAK and NCLAK groups. Statistical significance was defined as P-values below 0.05.

## 3. Result

The study comprised 60 patients with microbial keratitis that had been clinically identified. These were classified into two groups: CLAK (19 cases) and NCLAK (41 cases).

Demographic Profile: Out of 60 cases, 29 were female (43.3%) and 31 were male (51.7%). The age group most commonly affected was between 21–40 years. CLAK was more frequent among younger, urban contact lens users, while NCLAK was common in older, rural populations with a history of trauma or environmental exposure.

### 3.1 AGE-WISE DISTRIBUTION (CONTACT-LENS USERS)

Two isolates from the 51–60 age group, five from the 41–50 age group, nine from the 30–40 age group, and fourteen from the 18–29 age group made up the 30 samples.

**Table 1.** “Age-wise distribution (Contact lens users)”

| AGE | NO. OF ISOLATES | PERCENTAGE% |
| --- | --- | --- |
| 18–29 | 14 | 46.7% |
| 30–40 | 9 | 30.0% |
| 41–50 | 5 | 16.7% |
| 51–60 | 2 | 6.6% |
| **TOTAL** | **30** | **100%** |

**Chart 1.**
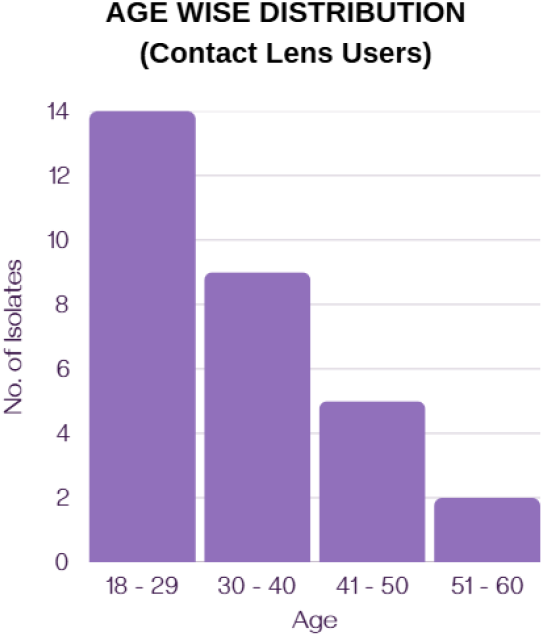
“Age-Wise Distribution of Contact-Lens Users”

### 3.2 “AGE-WISE DISTRIBUTION (NON-CONTACT LENS USERS)”

Out of 30 samples, 11 isolates were from the “18–29 age group”, 8 from the “30–40 age group”, 4 from the “41–50 age group”, and 7 from the “51–60 age group”.

**Table 2.** “Age-wise distribution (non-contact lens users)”

| AGE | NO. OF ISOLATES | PERCENTAGE% |
| --- | --- | --- |
| 18–29 | 11 | 36.7% |
| 30–40 | 8 | 26.7% |
| 41–50 | 4 | 13.3% |
| 51–60 | 7 | 23.3% |
| **TOTAL** | **30** | **100%** |

**Chart 2.**
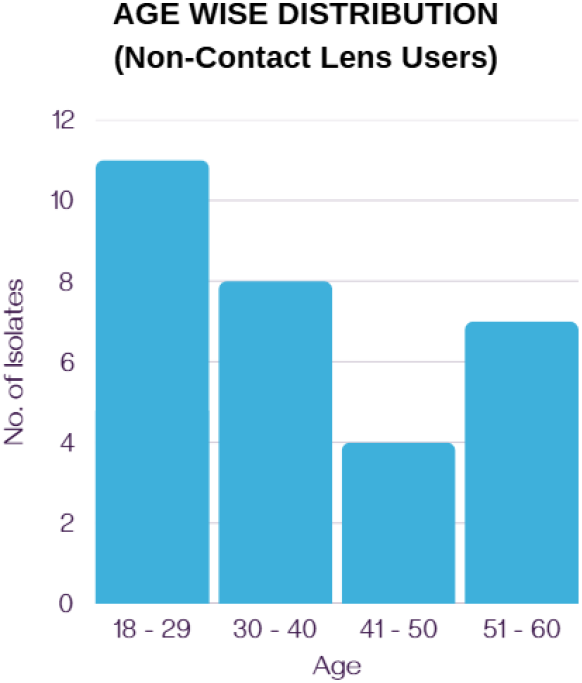
“Age-Wise Distribution of Non-Contact Lens Users”

### 3.3 GENDER WISE DISTRIBUTION IN CONTACT LENS ASSOCIATED KERATITIS (CLAK) AND NON-CONTACT LENS ASSOCIATED KERATITIS PATIENTS (NCLAK)

In a total of 29 females, 13 patients were Contact lens users, and 16 patients were Non-Contact lens users. Of 31 males, 6 patients were Contact lens users and 25 were Non-Contact lens users

**Chart 3.**
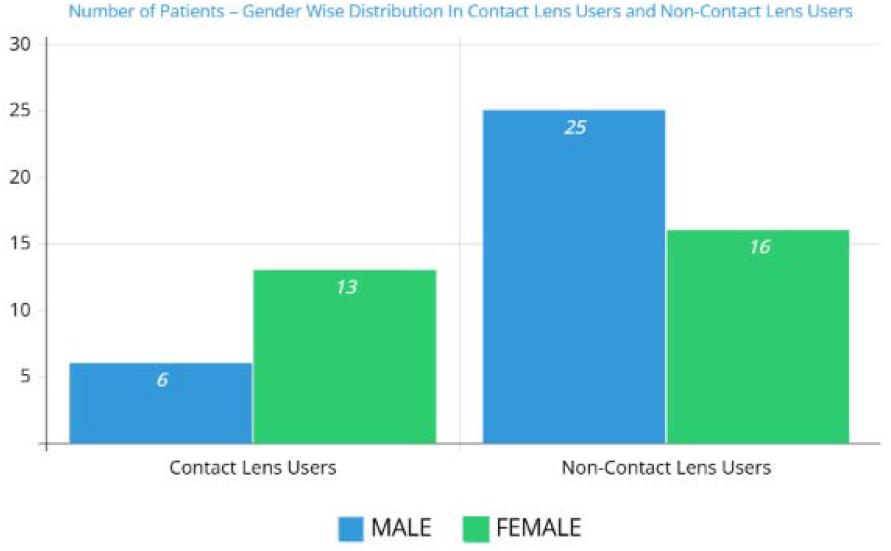
Number of Patients – Gender Wise Distribution in Contact Lens Users and Non-Contact Lens Users

**Table 3.** Gender wise distribution in CLAK and NCLAK.

| **SEX** | **CONTACT LENS**<br>**USERS** | **NON-CONTACT**<br>**LENS USERS** |
| --- | --- | --- |
| MALE | 6 | 25 |
| FEMALE | 13 | 16 |
| **TOTAL** | **19** | **41** |

### 3.4 OCCUPATIONAL DISTRIBUTION

Among all the patients, 30% of them were engaged in agriculture, 20% were manual labourers, and the remaining 50% comprised office workers, students, and homemakers.

**Table 4.** Occupational distribution of the patients.

| **OCCUPATION** | **NO. OF ISOLATES** |
| --- | --- |
| Office Workers,<br>Students, and<br>Homemakers | 50% |
| Agriculture | 30% |
| Manual Labourers | 20% |
| **TOTAL** | **100%** |

**Chart 4.**
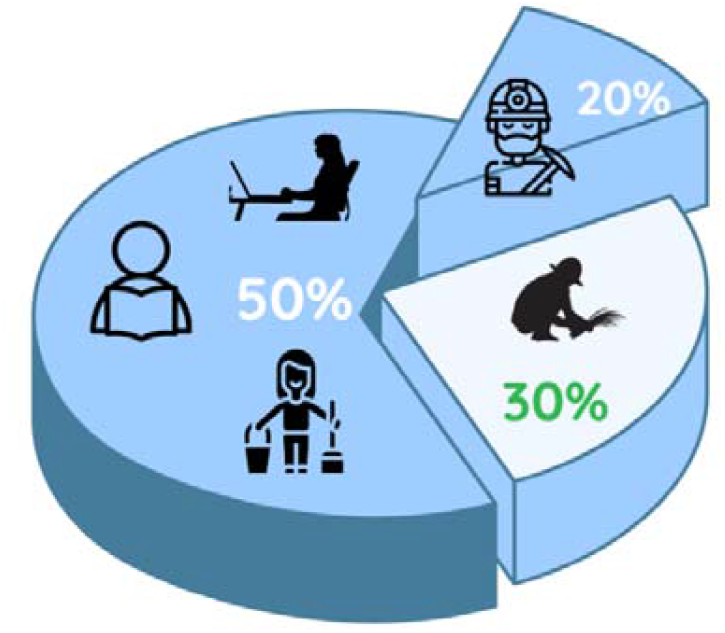
Showing Occupational Distribution of the patients

### 3.5 DISTRIBUTION OF MICROBIAL ISOLATES FROM DIFFERENT OCULAR SAMPLE TYPES IN KERATITIS PATIENTS

Out of 60 samples, 31 were collected from eye swabs, 4 from corneal scrapings, and 25 from contact lenses flushed in saline.

**Table 5.** Distribution of microbial isolates from different ocular sample types in keratitis patients.

| OCULAR ISOLATES | NO. OF SAMPLE | PERCENTAGE% |
| --- | --- | --- |
| Eye Swab | 31 | 51.7% |
| Corneal Scrapings | 4 | 6.7% |
| Contact Lens Flushed in Saline | 25 | 41.6% |
| **TOTAL** | **60** | **100%** |

**Venn Diagram 1.**
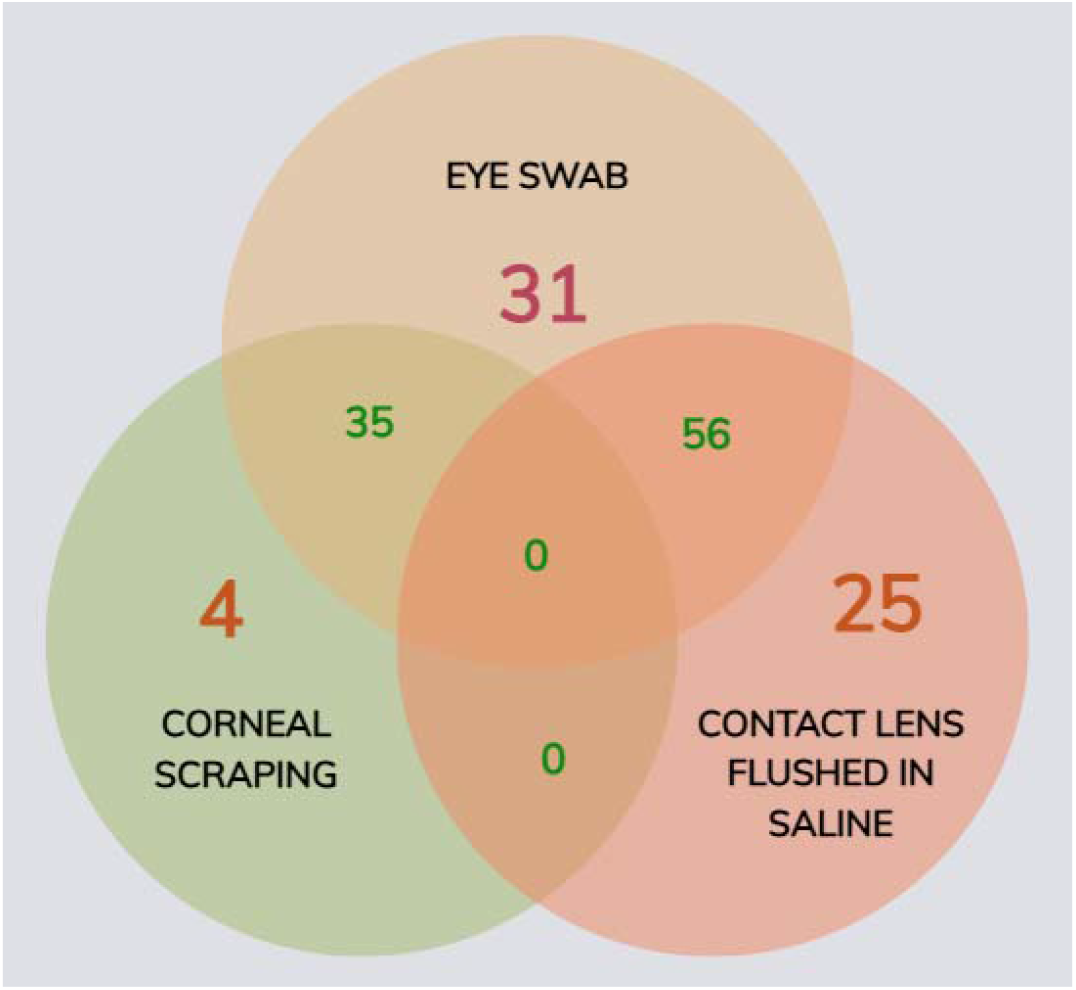
Showing the distribution of ocular isolates from different ocular sample types

### 3.6 ISOLATED BACTERIAL PATHOGENS

The isolated bacterial pathogens from the samples include *Pseudomonas aeruginosa* (16 isolates), *Escherichia coli* (10 isolates), *Staphylococcus aureus* (13 isolates), *Streptococcus pneumoniae* (8 isolates), and *Klebsiella pneumoniae* (6 isolates).

**Chart 6.**
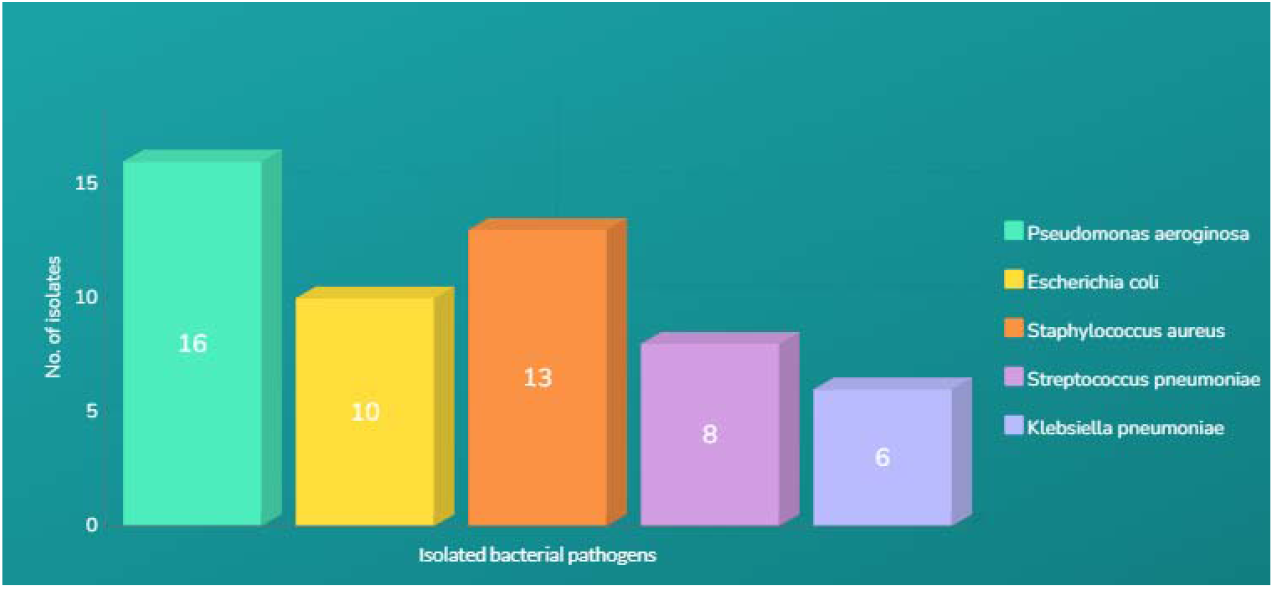
Bacterial Pathogens isolated from keratitis patients

**Table 6.** Isolated Bacterial pathogens from keratitis patients.

| S.NO | ISOLATED PATHOGENS | NO. OF ISOLATES |
| --- | --- | --- |
| 1 | *Escherichia coli* | 10 |
| 2 | *Staphylococcus aureus* | 13 |
| 3 | *Pseudomonas aeruginosa* | 16 |
| 4 | *Klebsiella pneumoniae* | 6 |
| 5 | *Streptococcus pneumoniae* | 8 |

### 3.7 ISOLATED FUNGAL PATHOGENS

The isolated fungal pathogens from the samples include Candida albicans (4 isolates), Aspergillus niger (2 isolates), and Aspergillus fumigatus (1 isolate).

**Table 7.**
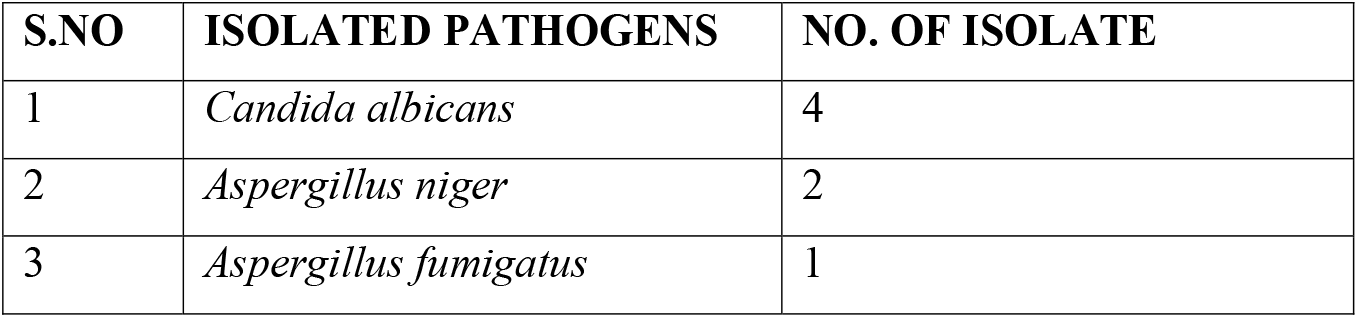
Isolated fungal pathogens from keratitis patients.

| S.NO | ISOLATED PATHOGENS | NO. OF ISOLATE |
| --- | --- | --- |
| 1 | *Candida albicans* | 4 |
| 2 | *Aspergillus niger* | 2 |
| 3 | *Aspergillus fumigatus* | 1 |

**Chart 7.**
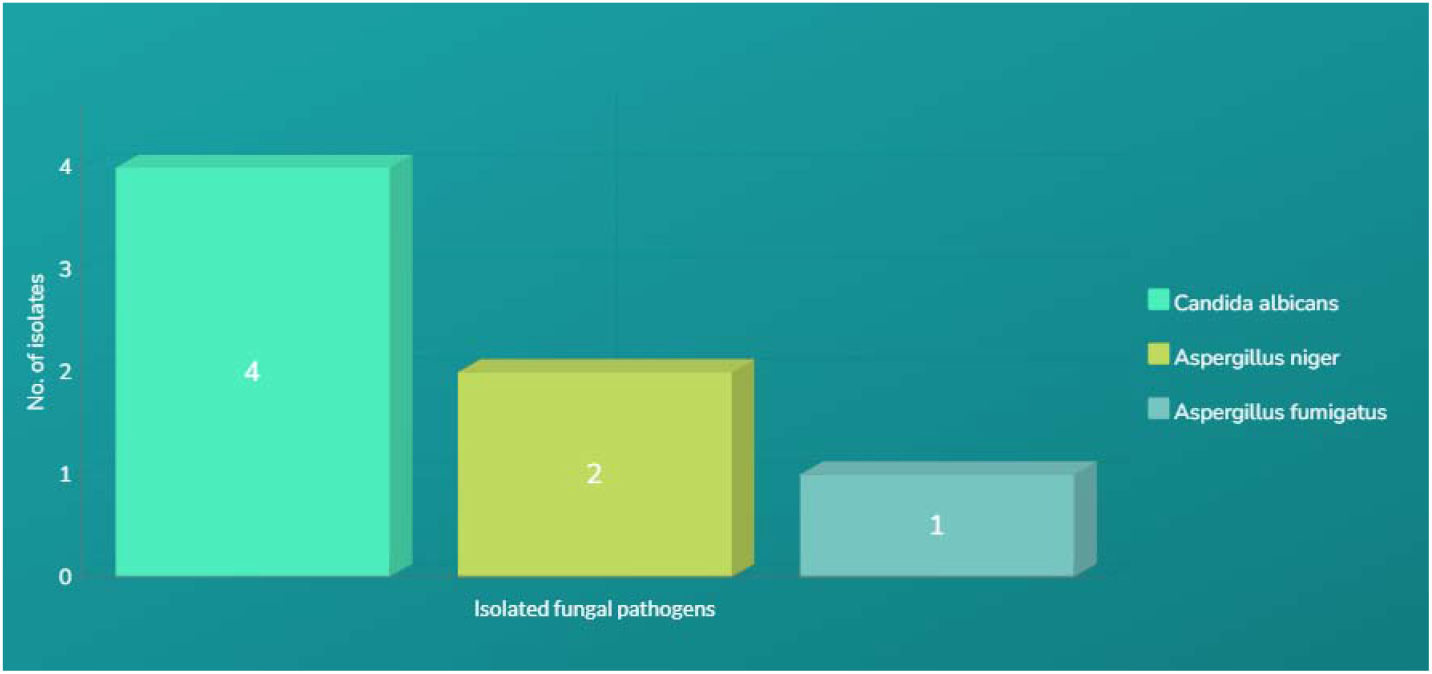
Fungal Pathogens isolated from keratitis patients

### 3.8 DISTRIBUTION AND GRAM STAINING CHARACTERISTICS OF BACTERIAL AND FUNGAL ISOLATES IN KERATITIS CASES

#### 3.8.1 MICROBIAL DISTRIBUTION IN KERATITIS

A total of 60 clinical samples were analysed to determine the presence of bacterial and fungal pathogens in keratitis cases. Among these, 25 samples (42%) contained a single bacterial species, predominantly *Pseudomonas aeruginosa, Staphylococcus aureus, Escherichia coli, Klebsiella pneumoniae*, and *Streptococcus pneumoniae*. Ten samples (17%) had co-infections with two bacterial species, with combinations such as *P. aeruginosa + S. aureus* and *E. coli +K. pneumoniae*.

**Table 8.** Distribution and Gram Staining characteristics of Bacterial and Fungal isolates in keratitis cases.

| Category | Microorganism | Gram Stain / Morphology | Number of Isolates |
| --- | --- | --- | --- |
| **Bacteria** | *Pseudomonas aeruginosa* | Gram-negative rod | 16 |
|  | *Staphylococcus aureus* | Gram-positive cocci (clusters) | 13 |
|  | *Escherichia coli* | Gram-negative rod | 10 |
|  | *Streptococcus pneumoniae* | Gram-positive cocci (lancet-shaped pairs) | 8 |
|  | *Klebsiella pneumoniae* | Gram-negative rod | 6 |
| **Fungi** | *Candida albicans* | Yeast cells with budding and pseudohyphae | 4 |
|  | *Aspergillus niger* | Filamentous fungus (septate hyphae) | 2 |
|  | *Aspergillus fumigatus* | Filamentous fungus (septate hyphae) | 1 |

Fungal infections alone were identified in 4 samples (12%), primarily caused by *Candida albicans, Aspergillus niger*, and *Aspergillus fumigatus*. However, a significant proportion of cases (20 samples, 33%) exhibited mixed infections involving both bacteria and fungi, suggesting a complex microbial interaction in keratitis pathogenesis. These mixed infections predominantly included bacterial species *(P. aeruginosa, S. aureus, E. coli, K. pneumoniae, S. pneumoniae)* in combination with fungal pathogens *(C. albicans, A. niger, and A. fumigatus)*.

**Table 9.** Microbial Distribution in Keratitis.

| Category | Number of Samples (n = 60) | Percentage (%) | Organisms Found |
| --- | --- | --- | --- |
| Single Bacteria | 25 | 42% | *Pseudomonas aeruginosa*, *Staphylococcus aureus*, *Escherichia coli*, *Klebsiella pneumoniae*, *Streptococcus pneumoniae* |
| Double Bacteria | 10 | 17% | Two of *P.aeruginosa*, *S.aureus*, *E.coli*, *K.pneumoniae*, *S.pneumoniae* (e.g., *P.aeruginosa* + *S.aureus*) |
| Only Fungi | 7 | 12% | *Candida albicans*, *Aspergillus niger*, *Aspergillus fumigatus* |
| Both Bacteria & Fungi | 18 | 30% | Combination of bacteria (*P.aeruginosa*, *S.aureus*, *E.coli*, *K.pneumoniae*, *S.pneumoniae*) + fungi (*C.albicans*, *A.niger*, *A.fumigatus*) |

**Chart 8.**
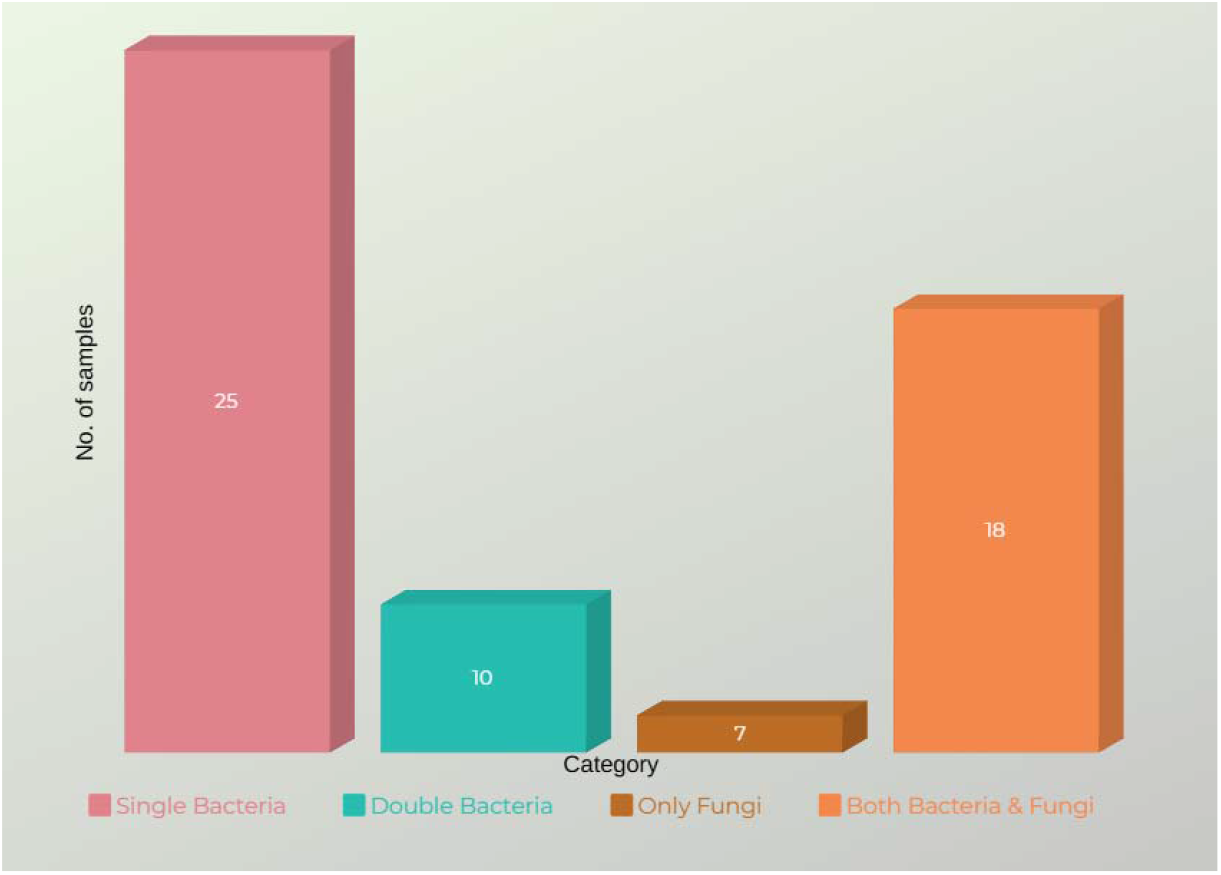
Microbial Distribution in Keratitis

### 3.9 BIOFILM ASSAY

**Figure 1.**
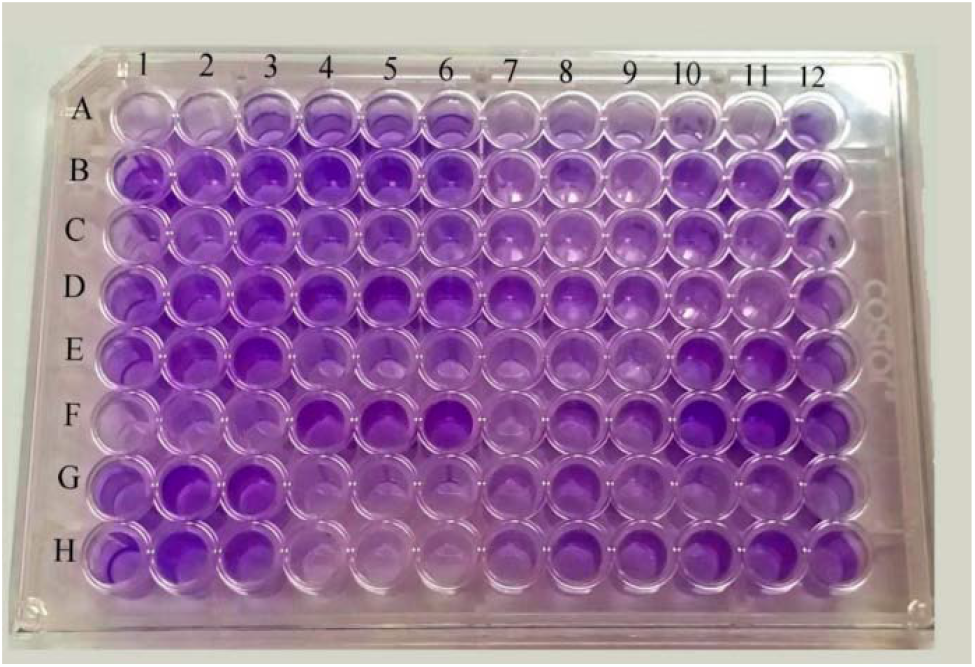
Biofilm Production Test Results “Biofilm production test by microtiter plate method: Negative Control (A1, A2, A3); Strong producer (F10, F11, F12); Moderate producer (B10, B11, B12); Weak producer (H7, H8, H9) and non-producer (H4, H5, H6). The higher the ODc, the higher the concentration of the purple color in the well”.

**Table 10.** Biofilm Production Results.

| S.NO | ORGANIMS | Biofilm Formation Strength |
| --- | --- | --- |
| 1 | *Escherichia coli* | Moderate |
| 2 | *Staphylococcus aureus* | Strong |
| 3 | *Pseudomonas aeruginosa* | Strong |
| 4 | *Klebsiella pneumoniae* | Moderate |
| 5 | *Streptococcus pneumoniae* | - |
| 6 | *Candida albicans* | Strong |
| 7 | *Aspergillus niger* | Weak |
| 8 | *Aspergillus fumigatus* | Weak |

## 4. Discussion

### Analysis of the Findings

In this investigation, bacterial and fungal isolates from contact-lens and non-contact lens-associated keratitis were examined for virulence factor. The findings offered a thorough comprehension of the pathogenic potential, and microbial genesis.

### Prevalence of Isolates

The bacterial isolates consisted principally of *Pseudomonas aeruginosa, Staphylococcus aureus, and Streptococcus pneumoniae*, while the fungal isolates included Aspergillus species and *Candida albicans*. Other investigations have found that *Pseudomonas aeruginosa* is a primary causal agent of microbiological keratitis, and it was substantially more prevalent in cases of keratitis linked to contact lens usage than in cases of keratitis unrelated to contact lens use, the studies of [13], [14] stated that *Pseudomonas aeruginosa* would adhere strongly to the corneal epithelium and it is also a strong biofilm producer.

Contact lens use and bacterial keratitis were significantly correlated (p < 0.05) according to a statistical study utilizing chi-square tests, which confirmed earlier epidemiological findings by Similar to findings in tropical countries, the increased prevalence of Candida in non-contact lens-associated keratitis cases points to environmental exposure as the main risk factor.

According to a breakdown of instances, fungal infections were more common in people who did not wear contact lenses, but bacterial infections predominated in people who did. This finding is in line with past epidemiological research that emphasize how warm, humid conditions might exacerbate fungal keratitis [15]. Additionally, P. aeruginosa strains showed clonal associations in the isolates’ genomic analysis, indicating a potential role for cross-contamination among lens users[16]

### Comparing This Study to Others

### The Distribution of Bacterial Pathogens

Studies of [17] [18] found a similar bacterial profile in keratitis cases, which is consistent with our data. Our results, however, indicated an increasing tendency, indicating a potential regional difference or shift in microbial dynamics, whereas their research reported a reduced prevalence of *Staphylococcus aureus*.

Our study also shows a notable presence of *Streptococcus pneumoniae*, which is frequently underrecognized as a keratitis cause.

### Restrictions and Prospects

Despite providing valuable insights, the study’s limitations include a small sample size and the lack of whole-genome sequencing to identify resistance determinants. Future research should focus on metagenomic methods and novel antimicrobial drugs to counteract resistance trends. Whole-genome sequencing might help detect the mutations that lead to antibiotic resistance, similar to the findings of the global surveillance research[19].

The results’ generalizability might also be improved by a more comprehensive epidemiological investigation that collected data from multiple centres and had a bigger sample size. As suggested by [20], investigating the host immune response to these infections may offer insights into novel therapeutic approaches.

## 5. Conclusion

This study highlights the significant differences in microbial etiology and biofilm production between contact lens-associated keratitis (CLAK) and non-contact lens-associated keratitis (NCLAK). Bacterial pathogens, particularly *Pseudomonas aeruginosa*, were predominantly isolated from CLAK cases, emphasizing the critical role of contact lens hygiene in preventing infection. In contrast, fungal pathogens, especially *Candida albicans* and *Aspergillus* species, were more common in NCLAK, often linked to environmental trauma. The strong biofilm-forming capacity of bacterial isolates, notably *Pseudomonas aeruginosa*, underscores the challenges in treatment due to increased antimicrobial resistance. Despite the fact that this study offers insightful information, the findings’ wider relevance is constrained by the small sample size and the absence of isolate molecular characterisation. To better understand pathogen behaviour and provide more effective prevention and treatment strategies for microbial keratitis, future research should concentrate on larger, multi-centric investigations, sophisticated genomic analyses, and investigation of host immune responses.

## Data Availability

All data produced in the present study are available upon reasonable request to the authors

## Authors Contribution

**Kavin Shilpa Abraham** conceptualized the study, performed a comprehensive literature review, synthesized the data, and prepared the initial and revised drafts. **Sujith Sri Surya Ravi** contributed to data interpretation and manuscript review, provided overall supervision, guided the scientific content, critically revised the manuscript, and approved the final version. **Leela Kagithakara Vajravelu** assisted in formatting, reference management, and final proofreading.

## Ethical Approval

Ethical approval was obtained from the Institutional Ethics Committee of **SRM MEDICAL COLLEGE HOSPITAL AND RESEARCH CENTRE** (Ethical Clearance Number: SRMIEC-ST0724-1400). Patient data and samples were anonymized prior to analysis to ensure confidentiality.

### Declaration of Generative AI and AI-assisted technologies in the writing process

The authors declare that generative artificial intelligence (AI) and AI-assisted technologies were not used in the writing process or any other process during the preparation of this manuscript.

## Acknowledgements

The authors would like to express their sincere gratitude to the Department of Microbiology, SRM Medical College Hospital and Research Centre, SRM Institute of Science and Technology, Kattankulathur, for providing the necessary academic support and resources throughout the preparation of this review. The authors also thank their peers and faculty members for their valuable insights and constructive feedback during the development of the manuscript.

